# A protocol for the multi-method evaluation of novel externally applied suction-based airway clearance devices in the treatment of foreign body airway obstructions

**DOI:** 10.1101/2021.09.11.21263431

**Authors:** Cody L Dunne, Ana Catarina Queiroga, David Szpilman, Kayla Viguers, Selena Osman, Amy E Peden

## Abstract

**Background:** Foreign body airway obstructions (FBAO, choking) are a significant cause of preventable mortality. Abdominal thrusts, back blows, and chest compressions are traditional interventions; however, suction-based airway clearance devices (anti-choking devices, ACDs) have recently been marketed as an alternative. Of note, there is limited published evidence regarding their efficacy and safety. Our research has two aims: 1) To investigate what situational and patient factors are frequently identified, and which are associated with relief of the FBAO, and survival, in individuals with a FBAO treated with an ACD; and 2) To describe the experience of individuals who have used ACDs in response to a FBAO and identify facilitators and barriers to the use of ACDs compared to traditional interventions.

**Methods and analysis:** All ACD case reports collected a priori by manufacturers will be analyzed up to July 1st, 2021. Following, a prospective database will be developed using an online reporting system to capture future ACD use from July 1st, 2021 to Dec 31st, 2023. Descriptive statistics will be used to summarize cases, 58 outcomes, and adverse events. Where possible, bivariable and multivariable analysis will be employed to assess for predictors of outcomes (relief of FBAO, survival, and survival with good neurological function). Semi-structured interviews will be conducted with a subset of ACD users to describe in detail their experience using the device. Themes from these interviews will be assessed using the Theoretical Domains Framework.

**Ethics and dissemination:** This study has ethics approval from the University of New South Wales Human Research Ethics Committee (HC210242). Findings from this multi-year, multi-method study will be published in peer reviewed literature, presented at conferences and contribute to informing future resuscitation guidelines. Data on ACDs are urgently needed as these devices are already being used by parents, caregivers, lay rescuers, and healthcare professionals to respond to choking emergencies.

**Strengths and limitations of the study:**

∘ Foreign body airway obstruction (FBAO) intervention is an area with limited data, largely based on retrospective case series for traditional techniques (e.g., abdominal thrusts, back blows and chest thrusts)
∘ A multi-phase, multi-year study will be conducted to evaluate a novel intervention (suction-based airway clearance devices, ACDs) that have the potential to improve pre-hospital response to foreign body airway obstructions and survival
∘ Utilization of retrospective and prospective data, as well as evaluation of the users’ experience, will help determine the role for airway clearance devices in resuscitation algorithms and future guidelines
∘ Data will be reliant on self-reporting from users due to the infrequent occurrence of FBAO and few ACDs available to general public
∘ Previous studies have relied on data collected by manufacturers, which will be improved here by providing users with an independent reporting method

## INTRODUCTION

Foreign body airway obstructions (FBAO, choking) are preventable injuries with a considerable mortality burden globally.[1-5] The outcome of a FBAO is determined largely by bystander actions taken outside of the hospital setting, and yet there is a paucity of contemporary data informing the best response to be undertaken by lay rescuers and first responders.[6]

The recent systematic review, authored by Couper et al., best summarizes the guiding literature in this area and highlights the limitations of current data.[6] Current recommendations from resuscitation councils, influenced by the International Liaison Committee on Resuscitation’s (ILCOR) Consensus on Science with Treatment Recommendations (CoSTR), suggest a combination of abdominal thrusts, back blows, and chest compressions as the core interventions for FBAO.[7] Table 1 summarizes the evidence currently available on which these recommendations are based. Of note, most data are restricted almost exclusively to case series, retrospective in nature, and no studies compare which intervention is superior in certain settings.

**Table 1.** Summary of evidence supporting current FBAO interventions from ILCOR’s most recent CoSTR [6-7].

| Intervention | Outcome | No. of studies and design(s) | No. of patients included |
| --- | --- | --- | --- |
| Abdominal Thrusts | Relief of FBAO | Six case series | 417 |
|  | Survival | Two case series | 189 |
| Back Blows | Relief of FBAO | Three case series | 75 |
|  | Survival | One case series | 13 |
| Chest Thrusts / Compressions | Relief of FBAO | One case series | 28 |
|  | Survival | One observational study | 138 (only 35 received the intervention) |
| Airway Clearance Devices | Relief of FBAO | One case series | 10 |
|  | Survival | One case series | 10 |

*FBAO = foreign body airway obstruction; No. = number;*

The limitations of this data reflect the challenges of conducting rigorous research in the field of resuscitation, especially in the pre-hospital environment. FBAOs are relatively rare events, and it is near impossible to conduct randomized studies where interventions are assigned as the vast majority are treated before Emergency Medical Service (EMS) workers arrive.[8] Further, reliance on only healthcare records (either from EMS or physicians) may omit nuances on situational factors or interventions that researchers are trying to evaluate. Conversely, data acquired from the memory of the patient or bystanders are subject to recall bias and are likely influenced by poor medical literacy.[9] This makes the data collected more limited and of lower quality compared to other areas of medical research. It also blurs the standard that new interventions must meet before being deemed safe and, subsequently, effective.

One new FBAO intervention that has been described in the past decade is airway clearance devices (anti-choking devices, ACDs). These externally applied, non-powered, portable suction devices have been marketed as an alternative when traditional FBAO techniques fail and fall into two broad types, that despite similarities, have considerable differences in their design. Non-invasive ACDs are primarily manufactured by LifeVac © (Fig 1A, LifeVac LLC, Nesconset, New York, USA), and consist of a facemask attached to a compressible bellows system.[10] Applying the mask to the choking person’s face creates a seal followed by a rapid depression of the plunger system and a firm pull upwards to deploy it. Minimally invasive ACDs are primarily manufactured by Dechoker © (Fig 1B, Dechoker LLC, Concord, North Carolina, USA), and consist of a short oropharyngeal tube that acts as a tongue depressor when the external facemask is applied.[11] The attached cylinder has a plunger that is pulled sharply to create the negative pressure system once a seal is formed with the mouth. Both contrast with other suction devices (e.g., Laerdal © V-Vac ®) that do not have the external anchor of the facemask and can enter deeper into the oral airway.[12]

**Fig 1.**
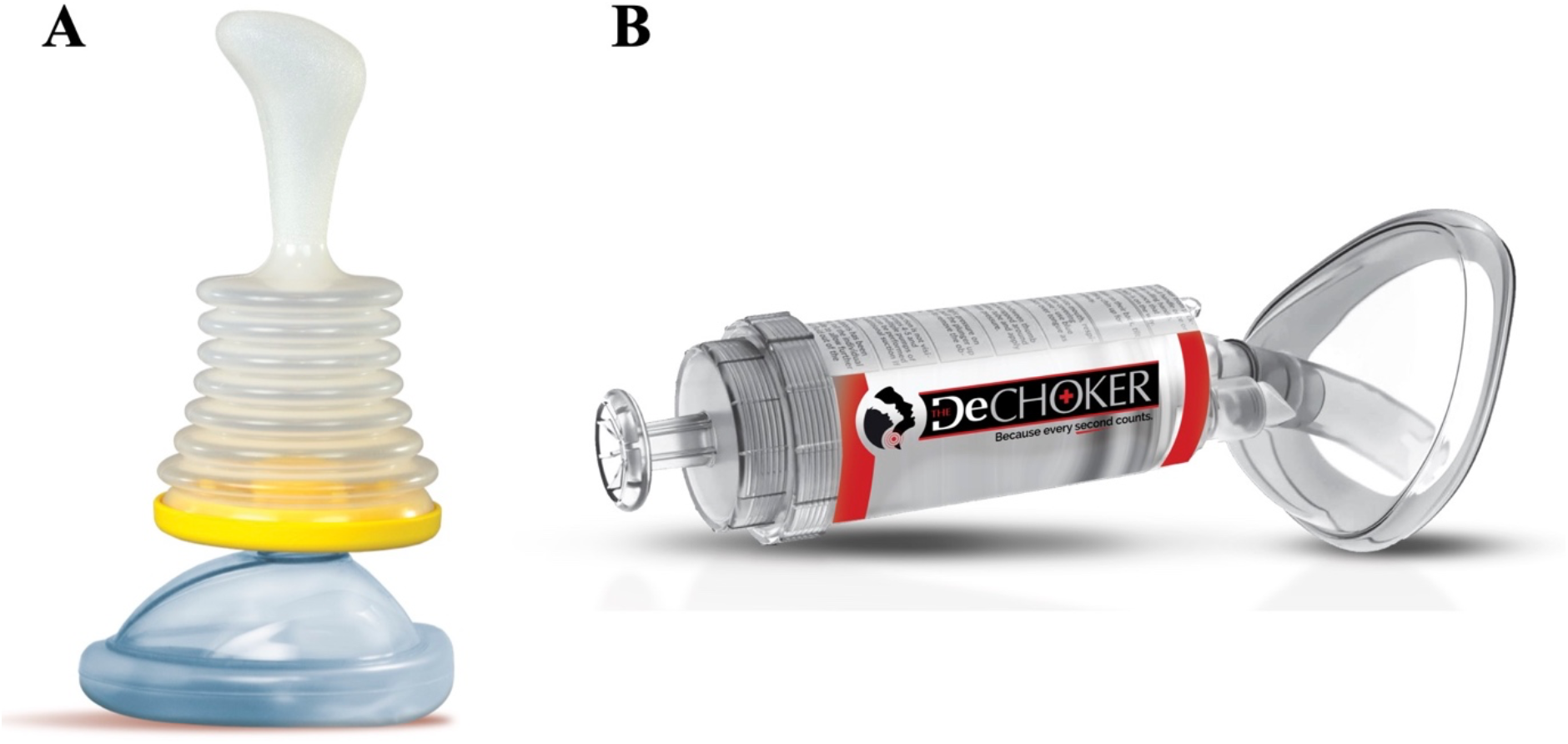
(A) LifeVac © airway clearance device (B) Dechoker © airway clearance device [images supplied by the respective manufacturers with permission to include].

However, two independent systematic reviews recently conducted concluded that there is insufficient evidence to support or discourage ACD use in the treatment of FBAO.[6, 13] Both reviews only identified ten unique cases describing the safety of LifeVac’s ACD (Table 1), all of which were successful at FBAO relief with no adverse events documented. They concluded with a recommendation that further evaluation in a research setting would be beneficial to determine the safety and effectiveness profile of ACDs.

Updated evidence in this area is required urgently. As sales of ACDs are increasing worldwide, the public and resuscitation councils require it to make informed decisions on their utility.[10, 11] If the safety and effectiveness profile remain promising, then acknowledging ACDs as an alternative to traditional techniques is appropriate. If not, then this also needs to be clarified. Regardless, the first step in this process is to conduct research, independent of industry, examining these outcomes further.

This protocol details the present study’s proposed method to address the following questions:

∘ <u>Research Aim One</u>: To investigate what situational and patient factors are frequently identified, and which are associated with relief of the FBAO, and survival, in individuals with a FBAO treated with an ACD.
∘ <u>Research Aim Two</u>: To describe the experience of individuals who have used ACDs in response to a FBAO and identify facilitators and barriers to the use of ACDs compared to traditional interventions.

## MATERIALS AND ANALYSIS

This is a multi-method, international observational study that will run from July 1^st^, 2021 – Dec 31^st^, 2023. Fig 2 depicts the project flow from design to publication. Three research studies will be used to address the two research aims.

**Fig 2.**
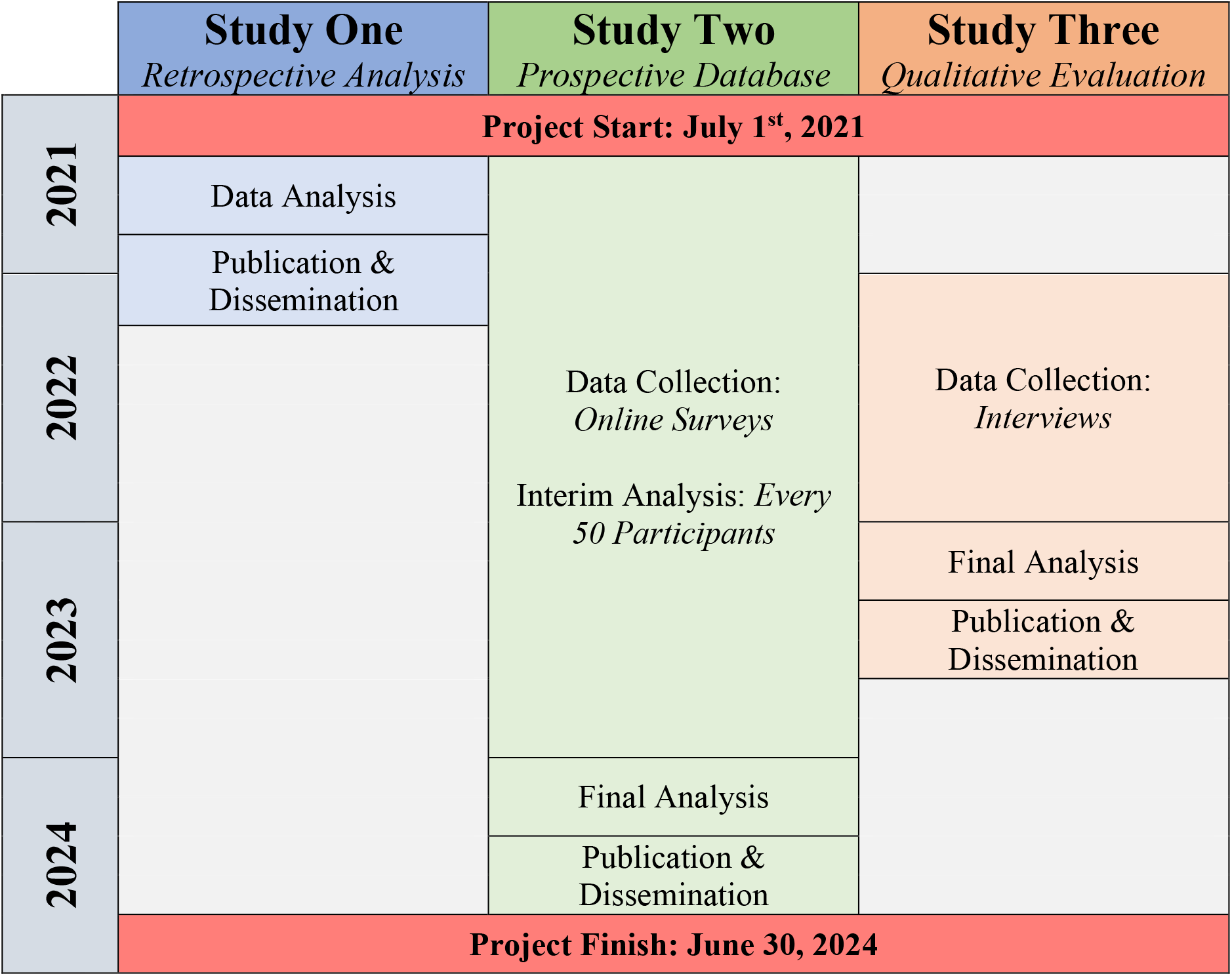
Visual depiction of the multi-year, multi-method evaluation of airway clearance devices.

### Study development

Following publication of the systematic review on the effectiveness of ACDs in January 2020, the research team met in late 2020 to discuss a strategy for future investigation.[13] A draft protocol was created and underwent several revision cycles until a final version was completed and agreed upon. In early 2021, contact was made with each manufacturer to arrange a meeting where an overview of the proposed study was introduced and discussed. The CEO of each respective company, as well as several members of their administration and/or medical team, were in attendance, along with the research team. Following this, the project protocol was provided to each company to review before confirming their interest in participating. In March 2021, letters of support were signed by the CEOs of both companies agreeing to participate to the extent that was outlined in the provided protocol, including assistance with identification and recruitment of participants, and providing access to their existing clinical report databases. The study was then reviewed and approved by the Heath Research Ethics Committee of University of New South Wales (HC210242) on May 25, 2021.

Following ethics approval, a subsequent meeting was coordinated with each company with a similar cohort of representatives as the initial meeting. Here, the protocol was reaffirmed by the research team and logistics were discussed regarding the transfer of anonymized data to the research team following July 1^st^, 2021 for the retrospective component and planning for the launch of the prospective component at this time. A member of the research team was available to answer any questions the manufacturers had regarding the study during this period via email or phone.

Of note, no significant changes to the study protocol were made after the initial meeting with the manufacturers.

### Study one: Retrospective evaluation of airway clearance devices

#### Participants and sample size

Participants will be all users of ACDs, who have self-identified before July 1^st^, 2021 to the device manufacturers as having experience utilizing ACDs on someone with a FBAO.

Sample size will be determined by the data collected a priori from the manufacturers. Based on reports from the manufacturers’ websites, this is estimated to be about 200 cases of ACD use in total.

#### Data collection

Both manufacturers have developed their own methods to allow customers who have used their ACD on a choking individual to report their experience. A waiver of consent for the secondary use of an existing dataset has been granted by the research ethics board for the use of the anonymized datasets.

Dechoker LLC began collecting data on its ACD’s real-life human uses in 2016. Dechoker’s reporting system varies based on its geographic region. In Europe and Australia, most sales are via local distributors and Dechoker relies on these smaller organizations to report back to them on any successful uses. In America, users contact Dechoker directly with their experience via electronic message or social media. Once identified, a member of the Dechoker team contacts the users to get further information on the situation and outcome.

LifeVac LLC began collecting data on its ACD’s real-life human uses in 2016. LifeVac has an online survey available on their website which participants can use to report their experience. This survey asks questions including the date of the incident, whether the Heimlich maneuver/back blows were performed, the consciousness of the patient at the time of ACD use, the FBAO’s severity and cause, and finally the outcome. Once a report is submitted, an administrative member of the company follows up with the user to confirm details and ask any further questions.

Both companies use social media extensively to promote their product, remind customers to report any uses to the company, and to identify events when ACDs have been used.

#### Data analysis

The data analysis will be similar to Study Two and is described in detail in the next section.

### Study two: Prospective database development and evaluation

#### Participants and recruitment procedure

Participants who meet the following criteria will be eligible for inclusion in the study:

1. Individuals 18 years of age or older, who have used an ACD on a human to attempt to dislodge a FBAO
2. Individuals who are not cognitively impaired, with no intellectual disability or a mental illness

The only exclusion criteria outside those not meeting the above will be anyone unwilling to provide consent, or those who are non-English or Spanish speaking (as the survey and interview guide will only be available in these languages).

ACD users will be invited to participate once they have been identified. Identification will occur in one of three methods. Both companies have agreed to include a location on their webpage where users can learn about the research study and access a link to the data collection survey on an independent site if they wish to participate. Companies also agreed to inform potential participants, who report an ACD use directly to them by email, phone, or social media, of the research study. A standardized e-mail was given to the companies to be sent to participants on behalf of the research team in this case. Finally, a website independent of all ACD manufacturers will be set up by the research team where interested individuals can learn more about the study and sign up to participate without involvement of the manufacturers. It is believed that this will empower users who have had an unfavourable experience with ACDs to feel comfortable reporting without the stress of doing so directly to the manufacturers.

The initial electronic message sent to all interested parties will contain a link to the online survey where participants will have access to the Participant Information Statement and Consent Form (PISCF) as well as the survey if they wish to proceed. At the end of the survey, participants will be able to indicate if they are interested in participating in an optional interview to help achieve Research Aim Two (see section below).

#### Sample size

There is no set sample size as this study is not designed to demonstrate superiority or non-inferiority to other FBAO techniques, and as such a power calculation is not applicable. Instead, the goal of this study is to increase the total data available in this field. The research team will conduct an interim analysis every 50 cases and decide to continue pending technical need for further data and availability of resources.

#### Data collection

Online surveys will be administered using digital survey software (Qualtrics, Provo, UT). The data collection tool was developed by the research team, and then beta-tested among ten individuals with no experience in healthcare or research. They were asked to provide feedback on the overall format of the tool as well as on any specific questions that were difficult to comprehend or were too complex to answer in a survey format. Based on their comments, the survey was revised prior to being uploaded to the Qualtrics system.

Data will be collected in four domains: participant (ACD user), patient (choking person), situational, and outcome (Table 2). Further, participants will be asked several agreement statements to further assess their experience.

**Table 2.** Summary of collected variables via online survey.

| **Data Category** | **Variables** |
| --- | --- |
| Participant (ACD user) | Demographics, relationship to person who choked, previous first aid or medical training, and previous experience with airway clearance devices (e.g., training, practice, or personal use) |
| Patient (Choking person) | Demographics, medical comorbidities, functional status pre-FBAO, and FBAO history |
| Situational | Date and location of FBAO, foreign body details, witnessed status, severity of FBAO, patient status at time of intervention (e.g., consciousness, crying/speaking), FBAO intervention details, ACD intervention details, CPR intervention details, and EMS activation |
| Outcome | Relief of FBAO, survival, survival with good neurological function, functional status post-FBAO, and adverse events |

*ACD = airway clearance device; CPR = cardiopulmonary resuscitation; EMS = emergency medical services; FBAO = foreign body airway obstruction;*

Participants will be provided with an email contact for the research team in case they are unsure how to respond to a particular question and want clarification before submitting their response. Further, all participants will be asked to consent to receive a follow up email in the case the research team requires clarification of their responses to ensure accuracy of responses, as well as confirmation the event occurred. Once their responses are checked by one of the research team members for clarity and follow up is conducted if needed, the results of the survey questions will be de-identified and assigned a unique study ID.

#### Data analysis

For both retrospective and prospective data, descriptive statistics will be used to summarize demographic, situational, outcome and adverse event data. This will include proportions, means [standard deviations], medians [interquartile ranges] with associated 95% confidence intervals as appropriate for the variable type.

Relief of FBAO by ACD will be defined as success/failure, survival as alive/dead, and survival with favourable neurological outcome will be assessed using the Cerebral Performance Category (CPC), with a favourable outcome being a category of 1 or 2 (good or moderate).[14] Functional status post-FBAO will also use the CPC. Adverse events will be summarized using descriptive statistics.

One of the medical researchers (SO, KV, CD) will assess information for each case to ensure appropriate assignment of the outcome. A subset of these (10%) will be checked randomly by one of the physician investigators (DS, CD) to ensure accuracy and reliability. Bivariable analysis will be performed using either the Student’s T (continuous) or Chi-squared test (categorical, Fisher’s Exact test if n<5) to measure the association of collected data with these outcomes. In all cases, statistical significance will be defined at the level of p < 0.05 for comparative analysis.

Where possible, adjusted multivariable analysis will be performed to determine which independent variables are able to discriminate between the main outcomes of clearance of the FBAO (i.e., success/failure), survival (alive/dead), and survival with good neurological outcome (CPC 1 or 2 / CPC 3, 4, or 5).

Where numbers permit, subgroup analysis will be performed for: adult versus pediatric patients, mild versus severe FBAO, patients with neurological and neurocognitive impairment, long term care residents, type of ACD used (e.g., LifeVac and Dechoker), and users’ prior training (e.g., first responder/healthcare worker versus first aid trained versus layperson without training).

If data are missing (or the input by the survey respondent is not usable), the proportion of missing data for each variable will be reported and for any analysis/calculations, the sample size (n) will be clearly identified.

### Study three: Qualitative evaluation of ACD users’ experiences

#### Participants and recruitment procedure

Participants in this study will be a subset of the individuals who participated in Study Two (Prospective Evaluation). Therefore, the same inclusion and exclusion criteria will apply, as will strategies for identification and recruitment of participants. Upon completion of the research survey, participants will be able to provide additional optional consent to be contacted by the research team to participate in an interview, further discussing their experience with ACDs.

Recruitment for the interviews will start on January 1^st^, 2022. A 6-month delay will occur to ensure the research design for the prospective study is functioning effectively before integrating the optional interview.

#### Sample size

Prior work utilizing the Theoretical Domains Framework (TDF) describes a stepwise approach.[15] Initially, 10 interviews will be conducted. After reaching that target, an analysis will be performed on the data collected to that point. Then three more interviews will be conducted, analyzed and the results compared to the original 10. If new themes emerged in the last three, another interview will be conducted. The stopping criteria will be defined as when three consecutive interviews produce no new themes compared to the prior (i.e., Interviews 1-11 will be compared to 12 – 14; then 1 – 12 to 13-15, and so on until no further novel themes are identified).

#### Data collection

Semi-structured interviews will be conducted using either online teleconferencing software or telephone. This will also allow data collection to be ongoing while respecting regional COVID-19 social distancing restrictions. An interview guide will be employed to facilitate the session and ensure consistency between interviewers. Interviews will be conducted by research team members trained on the interview guide. Each interviewer will have a rehearsal session with a simulated participant monitored by the primary investigator. Subsequently, they will all have their initial and randomly selected interviews monitored by another investigator to ensure a safe and high-quality experience for all involved. The interviews will be audio-recorded and then subsequently transcribed by the researchers using transcription software. A random selection of interviews (10%) will be manually transcribed by the research team, and the results compared to the transcription software to ensure accuracy in the results.

#### Data analysis

TDF will be used to guide the exploration of the ACD user’s experience and help identify facilitators and barriers to their use.[16] Qualitative data will be deductively analyzed, using TDF themes. Two independent researchers (CD, SO, KV) will code themes from interviews to domains of TDF and then compare for agreement. The coders will be blinded to the type of ACD used by the participant. Quantitative data will be reported as means [standard deviations] or medians [interquartile ranges] and for measures of association, the Student’s T or Chi-squared test will be used as appropriate. Confidence intervals will be reported, and statistical significance will be defined as p < 0.05. Agreement between coders will be measured using Cohen’s Kappa.

### Data management and privacy

All survey results and interview transcripts will be encrypted and stored on a secure server with two-factor authentication maintained by the University of New South Wales.

Retrospective data will be received from the device manufacturers de-identified. Prospective survey and interview data will be assigned a unique study ID and results will be separated from any identifying information specific to the participant. The code linking the unique study ID to the participant identification will be kept on a separate device stored separately from the data. The code will only be accessible to the chief investigator (AP) and delegate (CD) as needed.

Audio recordings will only contain the participants’ first name, but during the transcription process, it will be replaced with the participant’s assigned unique study ID to de-identify the record. Once transcribed and validated, the audio recordings will be destroyed.

No individual patient or healthcare provider information will be released at any point. The quantitative results will be presented in aggregate form to ensure the confidentiality of the participants. Direct quotations from interviews included in publications will not involve any identifying or case-specific information.

### Patient and public involvement

The public was involved in the development and piloting of the prospective survey which will be used to collect relevant, accurate data regarding ACDs. Critical changes were employed as a result of their involvement to improve purpose and clarity. All participants in the prospective evaluation will be given the opportunity to receive the results of the study via email. Further, the researchers will disseminate through conference presentations and develop a plain language summary of the result which will be provided to the manufacturers for promotion via social media interfaces.

### Study timeline

The study will be conducted according to the following timeline depicted in Table 3. This complements the information shown in Figure 1 above.

**Table 3.**
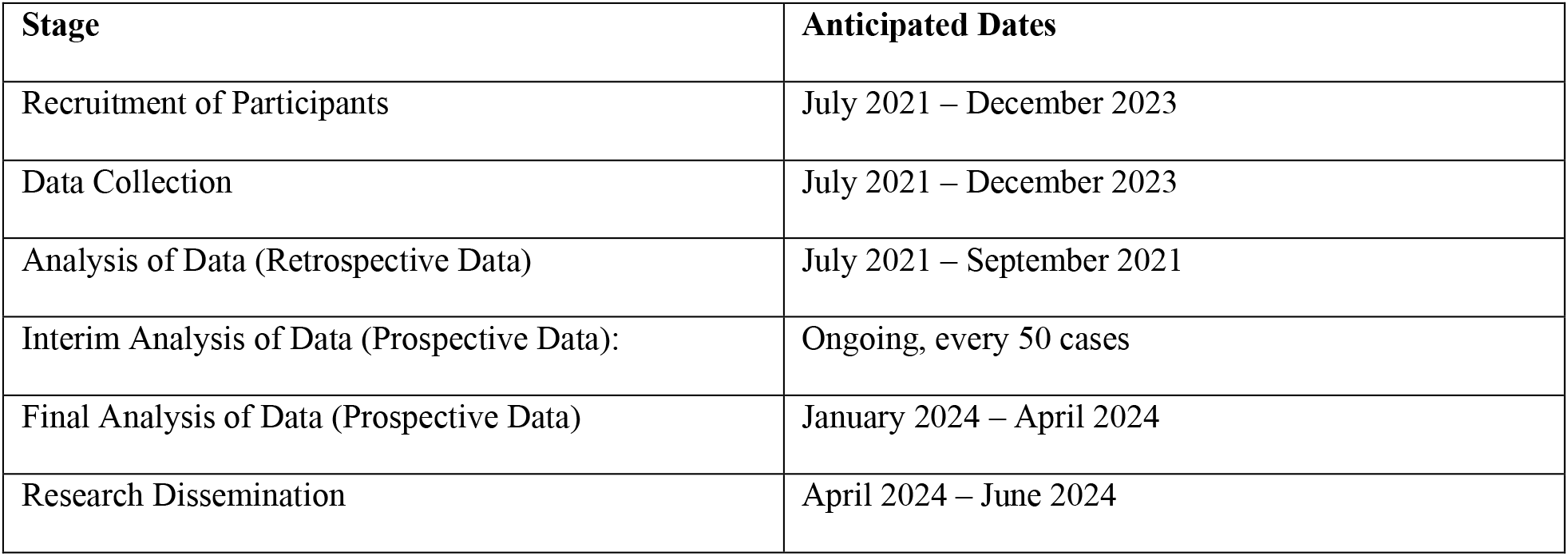
Anticipated research study timeline.

## ETHICS AND DISSEMINATION

This multi-year, multi-method study will evaluate clinical and safety outcomes of ACDs with the goal of increasing the available literature in this field to inform future resuscitation guidelines. It will employ a staggered design by first independently analyzing the retrospective secondary data previously made available by manufacturers, then create a prospective database of future cases. The latter component will be collected and analyzed independent of industry, with a protocol published a priori, clearly defined outcomes, and a publicly available reporting system.

### Strengths and limitations

This study will improve the quality of the evidence collected and inform future guidelines for use. Results of this study will impact global response to FBAO, and potentially save lives by improving the clearance rate of FBAO and stopping the progress of a choking person to cardiac arrest. By employing a staggered design, it will build on early findings from the retrospective analysis to identify key gaps in the data early and ensure improvement in the quality of data as the prospective component is phased in.

However, this study has several limitations. First, being observational in nature, any conclusions reached must be interpreted with caution as they are at increased risk of bias. However, the data in this field currently relies entirely on observational data and hopefully this will allow a comparison between existing and novel findings.[6]

Several classes of bias may be implicated in data gathering for this study. First, there is a high risk of confounding. As such, there may be different situations in which the ACD was used, and aggregation of all data may not represent true effectiveness in all circumstances. We will attempt to minimize this by being detailed in the variables collected, and if possible, conduct an adjusted analysis to minimize potential confounding. Still, there is potential that unmeasured variables may also be impacting the clinical outcomes. Second, as we are not randomizing users to interventions (i.e., ACDs or traditional techniques), we cannot control or completely adjust for the order they use an intervention or the number of attempts of each. Therefore, some users may go immediately to ACDs (despite statements that it is to be used only when other techniques have failed). This would likely increase the apparent effect as a shorter time to FBAO relief would be linked to improved outcomes. Whereas, others may do multiple traditional techniques before employing the device, delaying the ACD intervention. Time is a critical confounder for all resuscitation interventions, and resuscitation time bias is certainly a factor here. This concept acknowledges that the longer a person requires resuscitation, the more interventions they will receive, but consequently the worse outcomes they are likely to have due to a longer period of being in a critical state.[17] Although initially described for cardiac arrest, its theory is applicable here as well. As data is self-reported, time cannot be accurately measured to apply a time-based analysis, however, this may be a consideration for future studies.

The self-reporting design is a limitation of itself, as individuals may be prone to recall bias.[9] This will be due to both poor recall during a stressful event, and a variable ability of laypeople to accurately describe medical conditions and outcomes. This will be addressed by having all survey responses reviewed prior to de-identification, and if uncertainty in a response is noted, participants who consented to a follow-up email will be contacted by medical members of the research team. Unfortunately, it is not feasible in an international study to obtain permission to collect medical records from multiple health regions for a single patient at a time. However, this should also be a consideration for future studies if ACDs become highly utilized in a local region.

Self-reporting as a form of identification will result in an ascertainment bias, and skew results towards extremes of outcomes, which are more likely to be reported.[18] Ideally, all ACD uses would be captured by the database to model the true population and true effectiveness of the devices. However, the present reporting system is not rigorous enough to ensure this. This study will attempt to mitigate this effect by developing a reporting method outside the involvement of the manufacturers, where users who had unsuccessful or adverse outcomes still feel safe reporting the details.

Despite these limitations, the research team believes it is important to collect, analyze, and present these data to the public in as unbiased fashion as possible. In a field devoid of evidence, the first step to improving knowledge is to create a base on which to build future studies and further evaluations.

## CONCLUSION

This study describes a research protocol outlining the first multi-year, multi-method, global evaluation of novel suction-based airway clearance devices with the aim of improving the available data in this field, and ultimately help inform evidence-based guidelines in the future.

## Data Availability

No data is associated with this manuscript as it is a protocol.

## Authors’ contributions

Conceptualization: CD, AP, ACQ, DS; Methodology: CD, AP, ACQ, SO, KV, DS; Supervision: CD; Writing (Original Draft Preparation): CD; Writing (Review & Editing): CD, AP, KV, SO, ACQ, DS.

